# Deprivation and Segregation in Ovarian cancer survival among African American Women: a mediated analysis

**DOI:** 10.1101/2022.12.06.22283162

**Authors:** Andrew B. Lawson, Joanne Kim, Courtney Johnson, Theresa Hastert, Elisa V. Bandera, Anthony J. Alberg, Paul Terry, Maxwell Akonde, Hannah Mandle, Michele L. Cote, Melissa Bondy, Jeffrey Marks, Lauren Peres, Joellen Schildkraut, Edward S. Peters

## Abstract

**Background:** Deprivation and segregation indices are often examined as possible explanations for observed health disparities in population-based studies. In this study we assessed the role of recognized deprivation and segregation indices specifically as they affect survival in a cohort of self-identified Black women diagnosed with ovarian cancer who enrolled in the African American Cancer Epidemiology Study (AACES).

**Methods:** Mediation analysis was used to examine the direct and indirect effects between deprivation or segregation and overall survival, via a Bayesian structural equation model with Gibbs variable selection.

**Results:** The results suggest that high SES -related indices have an association with increased survival, ranging from 25%-56%. In contrast, ICE-race does not have a significant impact on overall survival. In many cases, the indirect effects have very wide credible intervals, consequently the total effect is not well estimated despite the estimation of the direct effect.

**Conclusions:** Our results show that black women living in higher SES neighborhoods are associated with increased survival with ovarian cancer, using area-level economic indices such as Yost or ICE-income. In addition, the KOLAK urbanization index has a similar impact, and the highlights importance of area-level deprivation and segregation as potentially modifiable social factors in ovarian cancer survival.

## INTRODUCTION

It is widely recognized that despite having a lower incidence of ovarian cancer black women experience poorer survival than white women. Although the reasons for this disparity are not clear, studies ^1^ have suggested that multiple social factors may play a role including socioeconomic factors. In recent years, a variety of area level deprivation indices (DIs) have been widely used in population-based epidemiologic studies as novel tools to examine the impact of neighborhood SES on a variety of health outcomes. Mechanisms by which neighborhood SES influences health could be indirectly through limited health care access or directly from cumulative biological effects of chronic stress. A recent extensive survey of the use of deprivation indices in cancer studies has been conducted.^2^ In that review, a number of commonly used indices (24) of deprivation were found to be associated with a range of cancer types in terms of incidence and survival or mortality. A few of these indices were most frequently used, such as the Yost, Concentrated Disadvantage Index (CDI), Area Deprivation Index (ADI), and Diez-Roux. In addition, the effects of structural racism and racial discrimination are increasingly being examined as a predictive factors in health disparities research. Segregation indices (SIs) can be formed based on race or SES and focus on the disparity between ethnicities or income at a local geographic level.

In the current study, we have sought to assess the effect of DIs and SIs on the survival of Black women diagnosed with ovarian cancer. In particular, we examined a cohort of Black epithelial ovarian cancer patients from the African American Cancer Epidemiology Study (AACES).^3^ It has been observed that contextual areal effects, such as the effects of residing in a particular locale or neighborhood, however, defined, can have an impact on cancer outcomes.^4^ In a previous analysis of patient characteristics (including histology), a disadvantage index measured at the census tract level was observed to be associated with ovarian cancer outcomes.^5^ In subsequent work,^6^ a disadvantage index at the census tract level was associated with poorer survival and later-stage outcomes in an ethnically mixed population. In addition, a significant relationship has been found between DI and ovarian cancer survival at the state and national levels.^7^ Previous studies in breast cancer, lung cancer, and prostate cancer have also found consistent relationships between disadvantage or deprivation and survival. ^8,9,10,11^ For this report, we have selected a set of DIs and SIs which represent different aspects of neighborhood SES context.

Our goal is to examine the association of differing attributes of DIs and SIs in relation to ovarian cancer survival metrics, including histologic subtype and diagnosis delays, and to consider the role of confounding and potential mediation in black women enrolled in AACES.

Our major hypotheses concerning DI and SIs are that these composite measures representing complex socio-economic status and social segregation neighborhood features have an important influence on the survival experience of women diagnosed with ovarian cancer whilst being mediated by important clinical and diagnostic factors.

To address these hypotheses, a suitable spatial scale must be assumed for the DI and Sis, which defines their context. We have chosen the census tract as that scale, and this is further discussed in the methods section below. In this paper, we pose the question: does living in a census tract with high deprivation or segregation lead to worsened survival from ovarian cancer in Black women?

## METHODS

### Study population

Phase 1 of the AACES study includes 11 US sites that were selected based on geographic regions with large African-American populations and to provide geographic diversity. These included Alabama, Georgia, Louisiana, metropolitan Detroit, Michigan, New Jersey, North Carolina, Ohio, South Carolina, Tennessee, Illinois, and Texas. Women diagnosed with epithelial ovarian cancer were eligible to participate if they self-identified as African American or Black, were 20-79 years of age at diagnosis, and could complete an interview in English.

Eligible cases had histologically confirmed epithelial ovarian cancer (EOC) and were centrally reviewed by an expert study pathologist. Rapid-case-ascertainment (RCA) was used to identify the cases through state cancer or Surveillance, Epidemiology, and End Results (SEER) registries, and through gynecologic oncology departments at individual hospitals. Participants were newly diagnosed with EOC between December 2010 and December 2015. The average and median survival time for all phase 1 cases, as of 2021, is 4.79 years and 4.81 years, respectively. The survival rates for this cohort are consistent with the observed survival rates from national SEER data,^12^ when conditioning on surviving at least 10 months past diagnosis. This accounts for the fact that rapidly fatal cases do not make it into the study, even when employing RCA. Approximately 50% of the phase 1 cases have participated in at least one follow-up survey. Altogether, 592 epithelial ovarian cancer cases were enrolled in phase 1. Of 1,720 cases attempted to contact, 1,199 (70%) were actively reached. Of these, 592 (49%) were successfully interviewed, and 388 (32%) actively refused. We are currently recruiting additional epithelial ovarian cancer cases in AACES phase 2, but these are excluded from the current analysis due to the brief follow-up time.

### Deprivation and Segregation indices

Deprivation indices are composite measures, usually based on socio-economic indicators. They are usually available within a range of small area units such as block groups (BGs), census tracts (CTs), or zip codes. In this study, we limit our focus to census tracts only, allowing for consistency across the available measures. Segregation measures address the division within an area between different population groups based on some key characteristics. These characteristics could be race-based or based on socio-economic status of the area. The following is a description of the chosen census tract DIs and SIs and their rationale for use.

#### Yost index

The Yost index has been constructed as an SES index via principal components analysis of seven variables.^13^ This index is widely used in cancer studies and is available at several geographic aggregation levels via the SEER database program (https://seer.cancer.gov/seerstat/databases/census-tract/index.html). This index was found to be significantly related to breast cancer incidence for a race stratified/adjusted analysis in the developers’ original study^13^ and other cancers in subsequent studies.^14^ A higher Yost value represents a higher SES level. We have used the index computed for a range of years: 2006– 2010 consistent with our case diagnosis years. The index is in standardized units.

#### Kolak urbanization (URB) measure

A recent study by Kolak of the social determinants of health^15^ proposed using four dimensions that encapsulate the variation in these determinants across spatial units within the US. The measures were derived from dimension reduction. The URB core dimension addresses urbanicity and reflects the degree to which an area has high rent, high income, and transport infrastructure and high costs for low-income residents. Unlike the Yost index, this measure does not directly reflect socio-economic status, but reflects a mix of factors related to urbanicity and hence urban and rural differences. The index is in standardized units.

#### ICE measures

The index of concentration at the extremes (ICE) are segregation measures (SIs) that can be computed at a given small area unit such as a block group or census tract.^16,17^ The indices estimate the differences in proportions of one group compared to an alternate group within an area relative to the total population, such as the proportion of the population that is Black compared to the White proportion. ICE-race is based on this comparison. ICE-income compares the lowest SES group in an area with the highest SES group. The calculation is based on the formula:

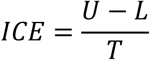

where *U* is the number of persons in the uppermost category, *L* is the number in the lowest, and *T* is the total population in the region in question. The ICE measures were computed from the 2010 census and the ACS 2006-2010. The indices range from -1 to +1 and a unit change represents a shift of 50% in the population structure.

#### The Spatial Context

In this study, we focus on a set of DIs as well as more recently developed segregation measures (SIs). It is important to consider the spatial level of resolution at which these indices are studied. The spatial context of deprivation is important as the influence of disadvantage on health could operate differently across various spatial scales. The EOC participants enrolled in phase 1 of AACES have been geo-coded to the census tract (CT) level. The social determinants of Health (SDOH) index (from now on Kolak indices), are only available at CT level. ^15^ For these reasons we have consider census tract measures as most relevant in our study.

AACES 1 consists of a sample of population-based registries. These registries are sparsely distributed across a selection of US states, and thus the spatial context is restricted. Our approach includes using observed individual predictors (such as age body mass index, physical activity self-reported SES) and also unobserved contextual effects within the spatial unit which allow for extra variation. In the modeling which follows we exploit a rich class of Bayesian hierarchical models (BHMs) which include predictors and random effects to allow for contextual confounding.

#### Mediation Methodology

To assess the impact of DIs and SIs on survival outcomes we apply a Bayesian causal mediation approach. ^18,19,20^ In this approach, a structural equation model is assumed whereby any mediator is modelled jointly with the outcome. In these joint models, the outcome is assumed to be a function of an exposure, a mediator, potential measured confounders, and a random effect that represents unobserved confounding. In this case, a Weibull end-point distribution is assumed where the log hazard is modelled with a linear predictor. In the typical mediator model, a dependence on exposure, confounders, and a random term representing unobserved extra variation is included. One of the significant advantages of Bayesian hierarchical models (BHMs) is the ability to incorporate unobserved variation via random effects.^21^ Note that the same random effect is shared and scaled in each of the outcome and mediator models. The mediation model structure ^22,23,2^ allows for the estimation of direct and indirect effects of predictors and we examine the paths within the directed dependence graph. The direct and indirect effects are estimated initially on the log scale and the relation between these is assumed to be *total effect = direct effect + mediator*mediator* parameter. The result is then exponentiated to give the odds estimate.

In the above model, a linear predictor for confounders is usually assumed to be fixed. However, we also consider an extension whereby we examine a range of putative models for confounders during our estimation procedure. A Bayesian method that allows the sampling of all models within the computational method of Markov chain Monte Carlo (MCMC) is Gibbs variable selection. ^24^ By allowing the sampling of different combinations of confounders, it is possible to estimate inclusion probability for any given variable. When the estimation process is complete, any confounder achieving an inclusion probability greater than 0.5 is selected for a final model. ^25^ The final model is fitted with only those confounders reaching this threshold. This avoids the use of stepwise selection methods and allows for all possible combinations of confounders to be assessed. The models are fit using the Nimble R package. ^26^ Additional details of the statistical approach can be found in the Appendix.

#### Mediation analysis Applied in AACES

##### Outcomes

We consider as our outcome the survival of the cohort participants. Here, survival is defined as the time in days from diagnosis to the date of death or last contact, updated through 2021.

##### Independent Variables

Our focus is on the effect of deprivation and segregation on EOC survival. As DIs or SIs are usually included as single adjustments in models, we assumed that, for our evaluation purposes, each index should be treated as the primary independent variable and fitted separately with mediators and confounders.

##### Mediators

Potential mediators were chosen *a priori* based on existing knowledge of their importance in EOC survival. The mediators included in this study were histology and diagnostic delay. Histology is regarded as an important prognostic indicator in EOC survival.^27^ Epithelial ovarian cancer histotype was determined through expert pathology review and was collapsed into high-grade serous or other for analysis. Additional mediators considered initially in this study included stage at diagnosis and residual disease (based on debulking status). Stage at diagnosis, is often used to reflect the severity of the disease.^27^ However, histology and stage are closely correlated and display similar mediation results, so we only discuss histology for brevity.

Diagnostic delay is included as a mediator as it could impact the severity of the diagnosed cancer and may be related to the socio-cultural environment of the locale or neighborhood. Diagnostic delay was defined in terms of the length of symptoms before the diagnosis. Ten symptoms were included in the baseline questionnaire. Specifically, participants were asked about symptoms in the year prior to diagnosis including pelvic or abdominal discomfort, changes in bowel movements, urinary issues, distended or hard abdomen, lump in the abdomen, fatigue, side or back pain, bleeding not related to menstruation, weight gain, and nausea. Diagnostic delay was summarized as the longest symptom duration before diagnosis, ranging from 1 to 12 months.

##### Confounders

We examined a range of both outcome and mediator confounders relevant in the analysis of survival outcomes: body mass index (BMI), smoking status, physical activity, age at diagnosis, and self-reported gross annual family income. BMI was calculated as kg/m^2^ from self-reported height and weight at a year prior to diagnosis. Smoking status (never smoker, former smoker, or current smoker) physical activity (amount of weekly physical activity (mild, moderate, and strenuous) reported in the year prior to diagnosis) was further categorized to capture inactivity (participants were considered inactive if their total weekly physical activity was less than 2 hours, and active if their total weekly physical activity was 2 hours or more), and family income (less than $10,000, $10,000-$24,999, $25,000-$49,999, $50,000-$74,999, $75,000-$100,000, and more than $100,000).

## RESULTS

Basic descriptive summaries of the selected clinical characteristics of the cohort appear in Table 1, and summaries of the DIs and Sis, and confounders are shown in Table 2. To frame the discussion, we note that significantly longer survival times are reflected as hazard ratios less than one. Any effect that significantly increases the hazard will show a ratio bounded above one. The original hazard is measured on the log scale, so the parameter estimates reported in Tables 3 and 4 have been exponentiated to yield odds. The estimates are reported with 95% credible intervals.

**Table 1.** Selected clinical characteristics of the cohort (N=558). The table count reflects the inclusion only of participants whose geocoding was complete.

| Characteristics | Mean (SD) |  |
| --- | --- | --- |
| Days | 1774.50 (962.86) |  |
| Delayed diagnosis (1-12 months)* | 6.98(4.34) |  |
|  | N | Percentage (%) |
| Survival Status |  |  |
| Alive (censored) | 221 | 39.6 |
| Deceased | 337 | 60.4 |
| Histology |  |  |
| High grade serous | 375 | 67.2 |
| Others | 183 | 32.8 |
| Stage |  |  |
| 1-Localized | 125 | 23.9 |
| 2-Regional | 51 | 9.8 |
| 3-Distant | 346 | 66.3 |
| Unknown | 36 | - |
| Debulking status – after imputation (residual disease) |  |  |
| 1=Optimal debulking (or CA125 after adjuvant <35) | 391 | 70.1 |
| 2=Suboptimal debulking (or CA125 after adjuvant ≥35) | 167 | 29.9 |

\*Delayed diagnosis variable has 64 missing observations

**Table 2.** Selected characteristics of the Dis, SIs and confounding variables. (N=558)

| Characteristics | Mean (SD) |  |
| --- | --- | --- |
| Age | 58.04(10.90) |  |
| BMI | 32.82(8.42) |  |
| Yost 2010 | 9493.66(921.62) |  |
| ICE race | -0.10(0.57) |  |
| ICE income | -0.19(0.27) |  |
| Kolak URB | -0.40(0.85) |  |
| Characteristics | N | Percentage (%) |
| Self-reported income |  |  |
| less than \$10,000 | 113 | 22.2 |
| \$10,000-\$24,999 | 120 | 23.6 |
| \$25,000-\$49,999 | 125 | 24.6 |
| \$50,000-\$74,999 | 76 | 14.9 |
| \$75,000-\$100,000 | 44 | 8.6 |
| More than \$100,000 | 31 | 6.1 |
| NA | 49 | - |
| Smoking status |  |  |
| Never smoker | 309 | 55.4 |
| Ever Smoker (Before/current) | 249 | 44.6 |
| PAGA |  |  |
| 1 = Yes | 130 | 25.1 |
| 2 = NO | 387 | 74.9 |
| NA | 41 | - |

\*Number of missing observations for the continuous variables
Kolak URB : 1, Yost: 13.

**Table 3:** Mediation analysis for ovarian cancer survival with the histology as the mediator variable. (Binary histology variable: 1. High grade Serous, 2. Others). The table reports the odds of increased (decreased) survival as influenced by the DI or SI reported. Well estimated odds shown in bold. The total effect is estimated from the direct effect and indirect effect estimated at each level of histology. In the final model the selected confounders were age alone (URB and Yost), and age and BMI (ICE indices).

|  | **URB** | **Yost** | **ICE-income** | **ICE - race** |
| --- | --- | --- | --- | --- |
| Direct Effect of each DI (each column) | **0.67 (0.51,0.849)** | **0.752 (0.593,0.946)** | **0.439 (0.193,0.922)** | 0.736 (0.509,1.049) |
| Indirect effect | 1.002 (0.635,1.5) | 1.014 (0.647,1.791) | 0.927 (0, 1.47x10<sup>14</sup>) | 0.988 (0.509,1.755) |
| Total effect | 0.671 (0.397,1.046) | 0.762 (0.437,1.415) | 0.407 (0,5.43x10<sup>13</sup>) | 0.727 (0.37,1.427) |
| Average histotype | 1.043 (0.61,1.951) | 0.991 (0.456,2.056) | 0.901 (0.366,1.896) | 0.714 (0.184,1.575) |
| Age | **1.05 (1.035,1.068)** | **1.051 (1.035,1.07)** | **1.055 (1.035,1.071)** | **1.043 (1.032,1.055)** |
| BMI | — | — | 1.208 (0.938,1.598) | 1.211 (0.958,1.562) |

**Table 4:** Mediation analysis for ovarian cancer survival with the delayed diagnosis of the cancer as the mediator variable. The table reports the odds of increased (decreased) survival as influenced by the DI or SI reported Well estimated odds shown in bold. In the final model the selected confounders were age and BMI for all the indices.

|  | URB | Yost | ICE-Income | ICE - race |
| --- | --- | --- | --- | --- |
| Direct Effect of each DI (each column) | **0.785**<br>**(0.678,0.904)** | **0.704**<br>**(0.542,0.937)** | **0.385**<br>**(0.124,0.901)** | 0.88<br>(0.718,1.074) |
| Indirect effect | 1.001<br>(0.994,1.009) | 1 (0.995,1.006) | 1.01<br>(0.972,1.057) | 1.007<br>(0.995,1.022) |
| Total effect | **0.785**<br>**(0.679,0.907)** | **0.704**<br>**(0.542,0.932)** | **0.389**<br>**(0.128,0.916)** | 0.886<br>(0.725,1.076) |
| Diagnostic delay | **0.900**<br>**(0.833,0.974)** | 0.995<br>(0.873,1.12) | **0.829**<br>**(0.729,0.926)** | **0.892**<br>**(0.826,0.961)** |
| Age | **1.032**<br>**(1.023,1.041)** | **0.929**<br>**(0.874,0.962)** | **1.078**<br>**(1.062,1.094)** | **1.032**<br>**(1.023,1.041)** |
| BMI | **1.175**<br>**(1.046,1.315)** | 0.773<br>(0.483,1.15) | **1.354**<br>**(1.067,1.717)** | **1.182**<br>**(1.052,1.34)** |

Table 3 displays the result of fitting models separately to the four indices as exposures with histology as mediator. The final model with selected confounders included only age for the URB and Yost indices, whereas the ICE indices had age and BMI. Notably, neither self-reported gross annual family income, smoking, nor physical activity are selected when histology is the mediator.

The major results in Table 3 illustrate that URB, Yost, and ICE-income are all associated with improved survival. ICE-race was also associated with improved survival but not statistically significant (OR = 0.736; 95% credible interval: 0.509-1.049). The results show that the Yost index, representing lower levels of deprivation, is associated with a 25% higher overall ovarian cancer survival per unit index (indirect effect: 1.014, 95% CI: 0.65, 1.79). The URB index, representing lower levels of deprivation and higher urbanicity, is associated with a 33% higher overall survival (indirect effect: 1.00, 95% CI: 0.64, 1.50). ICE-income, which represents higher levels of income segregation and a larger high income population, is associated with a 56% increase in ovarian cancer survival (indirect effect: 0.93, 95% CI: 0.0,1.47 × 10^14^). ICE-race, representing higher levels of segregation by race, showed no significant association with ovarian cancer survival. None of the indices we examined were medicated by histology when controlling for age and BMI. The indirect effects are also weakly estimated and centered on one, in some cases with wide credible intervals, thus total effects have wide intervals because of this. This suggests that histology has a limited effect as a mediator with the selected Dis and SIs. Note that the average effect of histotype is also not significant. In a separate analysis with the stage at diagnosis (data not shown), stage was also not found to be significant as a mediator. In summary, older age is associated with worse survival, whereas increases in the Dis and SIs are associated with longer survival, although the association for ICE-race was not statistically significant.

Table 4 displays the results of fitting models separately to the four indices as exposures with diagnostic delay as a mediator. Note that diagnostic delay is a continuous variable. In this case, age and BMI were selected for the final model for all indices. The direct and total estimated effects are more precisely estimated as the indirect effects have very narrow credible intervals, despite the indirect effects lacking statistical significance. URB, Yost, and ICE-income all have reduced odds and associated longer survival. ICE-race again reduces odds but does not achieve significance (OR = 0.88, 95% credible interval: 0.718-1.074). In addition, under this mediator, the ICE-income achieves the biggest reduction in survival compared with the other indices (0.385). However, a unit change in ICE represents a large population change (50%). Examination of the distribution of diagnostic delay times demonstrates that shorter delays are often associated with early mortality, and longer delays are found in the group with longer survival. Hence, this mediator displays a marginally enhanced effect on survival time. Similar interpretations for the four indices, as cited above in Table 3, can be made for the diagnostic delay mediator in Table 4.

## DISCUSSION

Our findings here suggest several important results for research on neighborhood effects on ovarian cancer survival. We observe that DIs and SIs do impact survival, although the degree of their effects differs. URB, Yost, and ICE-income all have a positive association with longer survival. This suggests that there is a ‘bootstrap’ (or health improvement) effect of living within a highly urbanized and high SES area with diverse income levels. The most marked direct effect observed is for ICE-income when a diagnostic delay is used as the mediator. However, this index also has an important impact when histotype is used as a mediator, demonstrating a nuanced relation between deprivation and survival, perhaps because early medical intervention often occurs when diagnosed at an advanced stage which is associated with worse survival even when the diagnosis is delayed.

In general, the different segregation indices differ in their association with survival. We observed that the SES-related index (ICE-income) displays a significant association with survival, with greater segregation income associated with improved survival. In contrast, the racial index (ICE-race) does not show any significance. This contrasts with other studies where ICE-race was observed to relate to racial disparity in an ethnically diverse population. ^27^ However, within our cohort, there was limited evidence of racial segregation (as measured by ICE-race) associated with survival. This could reflect the ethnic focus of the AACES cohort, while survival differences are often displayed when Black people are compared to White people in other studies. ^28^ This suggests that even concentrated wealth regardless of race has an effect on survival. The ranking of indices in terms of the greatest reduction in odds differs by a mediator. Yost and ICE-Income are lowest for diagnostic delay, and for histology, URB and ICE-income are lower.

Ross et al. ^14^ used the Yost index for SES to explore racial disparities in ovarian cancer survival and found that while SES may be associated with survival disparities by race, these disparities remained when adjusting for the Yost index. Hodeib et al.^29^ found that the Yost index was also a predictor of treatment guideline adherence for women with early-stage ovarian cancer. Our study shows that Yost SES at the census level has a significant direct effect overall effect on survival regardless of the mediator.

There are few additional studies investigating the role of other deprivation indices in ovarian cancer survival. An exception is Hufnagel et al. ^7^ where an increase in area deprivation index (ADI) was found to affect ovarian cancer survival negatively. A statistically significant association has been found between ovarian cancer-specific survival and Cagney’s concentrated disadvantage factor (CDI), ^5,30,^ which attenuates the disparity in survival between Black women and White women. A higher concentrated disadvantage was associated with poorer survival. Neither histology nor stage was found to have significant mediation effects with the chosen indices, perhaps because there is no effective screening protocol for ovarian cancer^31^.

Mediation effects differ according to the mediator we tested but generally do not add information to the relationship between these indices and survival. The direct effects dominate the total effect of the deprivation and segregation indices. Both histology and diagnostic delay have relatively narrow indirect credible intervals, and diagnostic delay has well-estimated total effects. Notably, ICE-income shows a considerable odds reduction of the hazard rate when mediated by diagnostic delay. This may suggest that a ‘bootstrap’ effect is in play, whereby differences in higher SES neighborhoods engender more prolonged survival. The URB index also shows a significant total effect with this mediator, which may reflect a similar bootstrap effect due to high urbanicity.

It should be noted however, that the mediation results cited are conditional on the presence of selected confounders. For example, while the inclusion of age and BMI with histology as a mediator led ICE-income to have much reduced odds (0.439), translating to a 56% increase in survival, the same confounders with diagnostic delay as a mediator led the ICE-income index to further improve survival (0.385), with a 62% increase in survival. Age shows a significant reduction in survival with histology, whereas it only shows a marginal improvement in survival under diagnostic delay. BMI is not selected under histology as a mediator, whereas with diagnostic delay, it is selected and shows a significant worsening of survival for URB and ICE-income. The importance of BMI and age in relation to the role of inflammation has been noted for this cohort. ^32^ A previous report using ICE-income/race was for breast cancer and found a significant association with the presence of ER+ status at the county level. ^16^ It is also notable that ICE-race are not found to be very well estimated under any mediator in our study. For ICE-income the biggest change was under diagnostic delay.

## Conclusions

This mediated analysis of DIs and SIs has wide-ranging implications for both neighborhood effects on ovarian cancer survival and the relative effectiveness of these indices when used in conjunction with different mediators and sets of confounders. First, commonly used indices such as Yost, ICE-income, and URB at the CT level, all significantly impact the survival experience of Black women diagnosed with epithelial ovarian cancer.

If the focus is to adjust for factors that could confound relations in ovarian cancer survival studies, then deprivation or segregation indices should be considered important general adjusters. In terms of confounders, age, BMI, or age alone were often selected within our variable selection strategy. However, self-reported SES, smoking, or physical activity did not appear as important confounders. Also, different mediators can affect the adjustment achieved by the use of DIs and SIs. While histology is important in explaining survival variation, ^28^ the different mediators behaved differently. The indices most strongly associated with survival in our analyses are Yost and ICE-income when mediated by diagnostic delay and URB and ICE-income when histology is present. ICE-race does not provide significant adjustment in survival.

There are a number of caveats to these conclusions which should be stressed. First, all analyses were based on census tract-level measures, so results could vary if other spatial units, such as block group or county, were chosen. Our cohort is purely self-identified Black women with ovarian cancer, and so disparities between races are not our focus, although there are results reported previously that found the ethnic disparity in survival for ovarian cancer was significantly associated with neighborhood disadvantage.^30^ Our mediated analysis did not examine multiple mediation as we confined our focus to single mediators. While multiple mediation is certainly feasible within our strategy, our focus on evaluating DIs and SIs limited our approach. In addition, our results are conditional on the inclusion of selected important confounders (mainly age and BMI) but also included random effects that allowed for unobserved confounding. Any analysis that did not adjust for these predictors would likely have extra noise. The inclusion of random effects is therefore crucial in allowing for this additional variation.

Finally, the most important outcomes are in validating contextual indices and their essential role in affecting survival among Black women diagnosed with ovarian cancer. The census tract appears to yield significant survival associations for both DIs and SIs. We have confirmed that the much used Yost index does indeed provide a validated indicator for extended survival, as does the Kolak urbanization dimension. The ICE indices also provide important stratification associations. These results attest to the utility of the Bayesian mediation strategy used and also to the relevance of the census tract as an important contextual spatial unit.

## Data Availability

Data used in this study is protected health information and could be made available if IRB approval is sought for the applicant.

### Appendix

In this Appendix the detail of the Bayesian joint mediation models is provided.

For a single mediator the following describes the model assumed: for the *i* th participant: the survival time is *T*_*i*_, and the censoring indicator is *δ*_*i*_. The mediator is defined to be *M*_*i*_, the exposure is *X*^*i*^, and the confounder set is {*C*_*i*_}. In addition, we assume that a random effect at the participant level will be included in the models: *U*_*i*_. For a survival outcome, we assume a conventional Weibull model, conditional on the mediator, exposure and observed and unobserved confounding, suppressing the individual subscript for convenience:

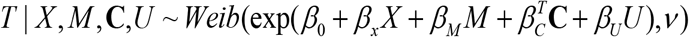

This choice of model allows for a flexible form of hazard. In this formulation, the term 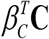 is a linear predictor as a function of the confounder set.

For the mediation model, conditional on exposure and confounders :

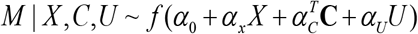

where *f* (.) is a chosen mediator distribution. Note that the same random effect is shared and scaled in each of the outcome and mediator models.

Finally, the random effect model, which, in general, can depend on exposure and confounders, is

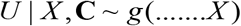

where *g* (.) is a suitable random effect distribution. This will be assumed to be continuous and zero mean Gaussian unless otherwise stated, and in our study will not be directly dependent on exposure or confounding. The above models are fitted jointly so that the random effect is shared but scaled. Note that the confounders are assumed to have different linear predictors in the outcome and mediator models.

In this model structure, the direct and indirect effects of an exposure is defined as: Direct effect: ≈ *β*_*x*_, Indirect effect: ≈α_*X*_ β_*M*_ and Total effect: β_*X*_ + α_*X*_ β_*M*_.

#### Variable Selection

In the above model, the linear predictor for confounders is assumed to be fixed. However, it is useful to consider an extension whereby we examine a range of putative models during our estimation procedure. A Bayesian method which allows the sampling of all models within the computational method of MCMC is Gibbs variable selection. ^23^

Within the design matrix **C** we can have multiple predictors and may wish to perform variable selection on these. Hence, we could replace 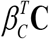 and 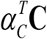 with 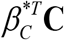 where 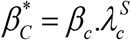 and 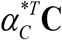 where 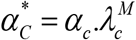 respectively. The λ parameters are inclusion or entry parameters and allow the predictors to be swapped in and out of the model during estimation. It is assumed that the ‘entry’ parameters have Bernoulli prior distributions 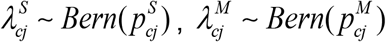:. This allows the inclusion parameters to range between [0,1]. When the estimation process is complete, the inclusion parameters are averaged and any predictor where *ave*(λ) > 0.5 is selected for a final model. ^24^ The final model is then fitted with only those predictors reaching this threshold.

#### Computation and Prior Distributions

To fit the above model, and all the model variants we discuss here, we have implemented the models using a MCMC procedure in the R package Nimble.^25^ In general, we assume regression parameters to have conventional zero mean Gaussian prior distributions with precisions which have *Ga(a,b)* prior distributions. We assume weaky informative gamma priors with a = 2.0 and b = 0.5.

Nimble chooses the most appropriate sampling algorithms to use to generate samples from the posterior distribution.

